# Application of Discriminant Analysis for Blood Pressure Classification Based on Vital Signs: Evidence from a Regional Hospital in Ghana

**DOI:** 10.64898/2026.03.06.26347774

**Authors:** Daniel Cobbinah, John Awuah Addor, Kingsley Macchrist Addo Narh, Emmanuel Mensah Baah

## Abstract

**Background and Aims:** Hypertension and diabetes mellitus are major non-communicable diseases and leading contributors to cardiovascular morbidity and mortality, particularly in low and middle-income countries. Both conditions frequently coexist and share common risk factors, including obesity and advancing age. Early identification of abnormal blood pressure status using routinely collected clinical data may enhance timely intervention and reduce complications associated with hypertension and diabetes. This study aimed to develop and validate a discriminant model that classifies patients as hypotensive, normotensive, or hypertensive using common vital sign indicators, and to evaluate the predictive contribution of individual variables within the broader context of cardio metabolic risk.

**Methods:** This retrospective observational study analyzed secondary data from 1,000 adult patients at a regional hospital in Ghana. Linear discriminant analysis (LDA) was applied using age, heart rate, body temperature, and body weight as predictors of systolic blood pressure classification. Model performance was assessed using cross-validation and classification matrices. Receiver operating characteristic (ROC) analysis was conducted to evaluate the discriminatory ability of individual predictors.

**Conclusions:** Routinely collected vital sign data, particularly body weight, can accurately classify blood pressure status. The high classification accuracy observed supports the feasibility of data-driven risk stratification in clinical settings. These findings further underscore the importance of weight management in the prevention and control of hypertension and related cardio metabolic conditions, including diabetes.

## 1. INTRODUCTION

Blood pressure regulation is a critical physiological process, and abnormal blood pressure levels particularly hypertension remain a major global public health concern. Hypertension is a leading risk factor for cardiovascular disease, stroke, renal failure, and premature mortality worldwide (WHO, 2013; Chobanian et al., 2003). In low- and middle-income countries, including Ghana, the burden of hypertension is rising due to demographic transitions, lifestyle changes, and limited access to early detection and preventive care.

Blood pressure is commonly classified into hypotensive, normotensive, and hypertensive states based on systolic and diastolic measurements. While direct blood pressure measurement is standard clinical practice, identifying individuals at risk using routinely collected vital indicators can enhance early screening, risk stratification, and targeted intervention, especially in resource-constrained health systems.

Existing literature demonstrates that several physiological and demographic factors influence blood pressure. Age is a well-established determinant, with blood pressure generally increasing as arterial stiffness and vascular resistance rise over the life course (Dorans et al., 2018; Wang & Wang, 2004). Body weight and adiposity are strongly associated with hypertension, with excess weight contributing to increased cardiac output and vascular resistance (Janssen et al., 2004; Neter et al., 2003; Hall et al., 2015). Temperature variation has also been linked to blood pressure regulation through thermoregulatory and vascular mechanisms. Studies show that cold exposure and seasonal temperature changes can influence systolic and diastolic pressure (Cabrera et al., 2016; Lewington et al., 2012; Umishio et al., 2019). Resting heart rate has been associated with future hypertension risk and cardiovascular stress (American Heart Association, 2023). Gender differences in hypertension prevalence and complications have also been reported, although patterns vary across populations (Chen et al., 2019; Modesti et al., 2006).

Despite these known associations, many empirical studies focus on isolated predictors or employ regression-based approaches that model blood pressure as a continuous outcome (Chen & Wang, 2008; Wang et al., 2022). Such approaches do not explicitly address the clinical task of classifying individuals into meaningful blood pressure categories. Moreover, limited attention has been given to multivariate classification techniques that assess the combined discriminatory power of routine vital signs in distinguishing hypotensive, normotensive, and hypertensive patients.

Discriminant analysis provides a robust multivariate framework for classifying individuals into predefined groups based on multiple predictors simultaneously. By maximizing between-group separation while minimizing within-group variability, this approach is well suited for clinical classification problems. Applying discriminant analysis to routinely collected hospital data can generate practical, interpretable models that support decision-making and population-level screening.

Conceptually, as can be seen from **Fig1** this study is grounded in the premise that vital indicators specifically age, body weight, body temperature, and heart rate are interrelated physiological markers that collectively influence blood pressure regulation. These indicators act as independent variables that contribute to variation in systolic blood pressure status, resulting in classification into hypotensive, normotensive, or hypertensive categories.

**Figure 1:**
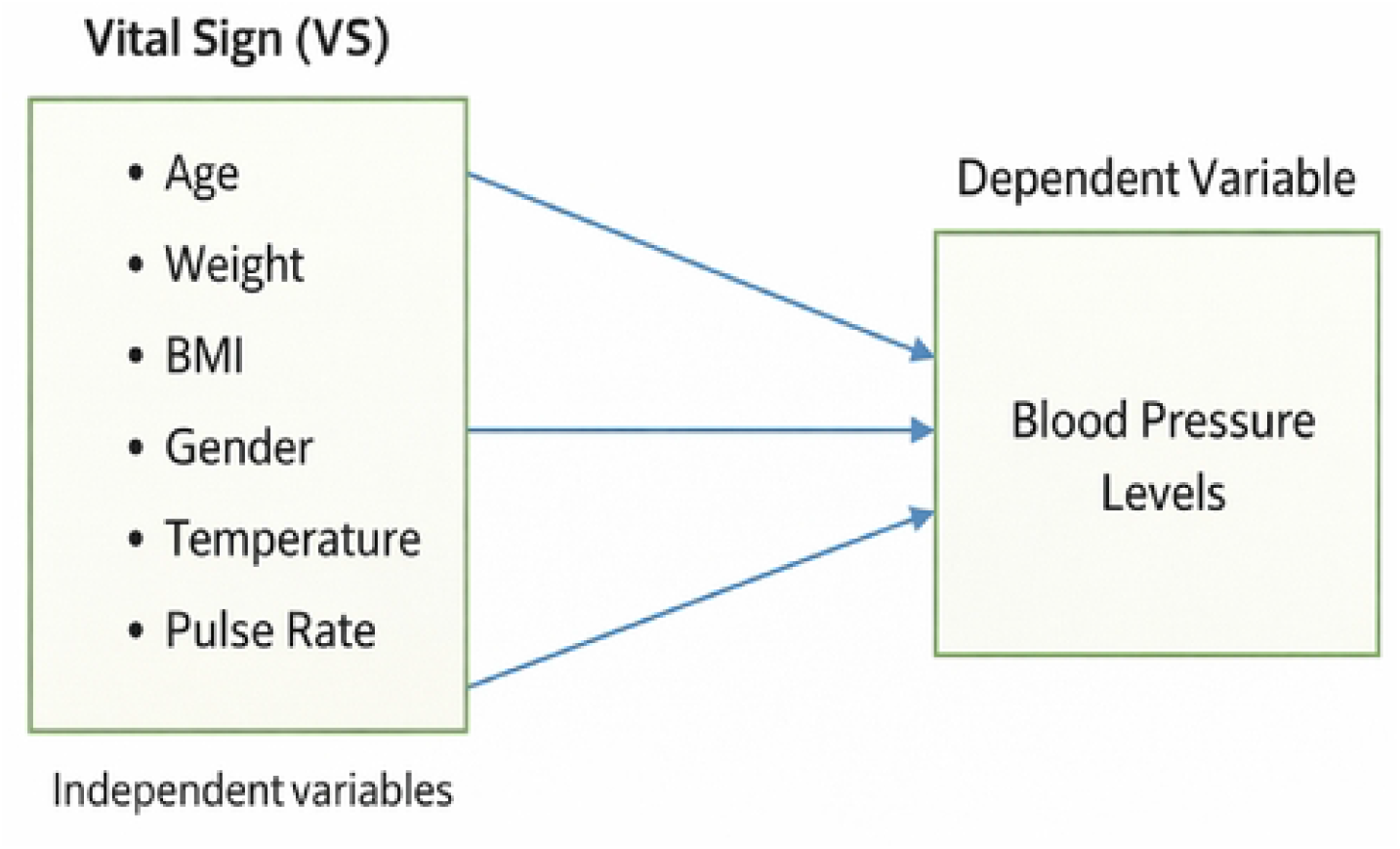
Discriminating Blood Pressure Levels Using Vital Indicators.

Therefore, the aim of this study is to develop and evaluate a discriminant model for classifying systolic blood pressure levels using routine vital indicators from hospital records in Ghana. The specific objectives are to:

1. Identify the discriminant model that best separates hypotensive, normotensive, and hypertensive patients
2. Assess the predictive performance and classification accuracy of the model
3. Evaluate the sensitivity and specificity of individual vital indicators using receiver operating characteristic (ROC) analysis

By leveraging routinely collected clinical data, this study contributes evidence supporting efficient and scalable approaches to blood pressure risk stratification in clinical and public health settings.

## 2. METHODS

This retrospective observational study used anonymized secondary data obtained from a regional hospital in Ghana following institutional permission for academic analysis and this was obtained on the 12 August, 2025. The dataset comprised records of 1,000 adult patients aged 18 years and above. Patients were categorized according to systolic blood pressure measurements into three clinical groups: hypotensive (< 90 mmHg), normotensive (90–120 mmHg), and hypertensive (> 120 mmHg). Predictor variables extracted from routine clinical records included age (years), body weight (kg), body temperature (°C), and heart rate (beats per minute). Linear discriminant analysis (LDA) was applied to classify patients into blood pressure categories using these physiological indicators. Key assumptions underlying LDA including multivariate normality, equality of covariance matrices, and absence of multicollinearity were assessed using standard diagnostic procedures. Variance inflation factor (VIF) statistics were used to verify that predictor variables did not exhibit problematic collinearity. Model performance was evaluated through classification matrices and cross-validation to assess predictive stability. Receiver operating characteristic (ROC) curves and corresponding area under the curve (AUC) statistics were computed to determine the discrimination ability of individual predictors. All statistical analyses were conducted using STATA and Python.

## 3. RESULT

### 3.1. Preliminary analysis

This section summarizes descriptive statistics of patient characteristics and evaluates assumptions required for Linear Discriminant Analysis (LDA).

#### 3.1.1 Basic statistics on patients’ vital indicators

**Table 1** presents the distribution of blood pressure categories by gender among 1,000 patients. Hypertension was the most prevalent condition (51%), followed by normotension (44%) and hypotension (5%). The distribution of blood pressure categories was similar across genders, indicating no marked gender-based differences in classification.

**Table 2** summarizes descriptive statistics for age, heart rate, body temperature, and weight. The mean age of patients was 45 years, with moderate variability across the sample. Heart rate and body temperature showed relatively narrow distributions, while body weight exhibited broader variation. Skewness analysis indicated that most variables approximated normal distributions, with temperature showing slight right skew due to a small number of outliers.

#### 3.2.1 Assumptions of Linear Discriminant Analysis

LDA assumes multivariate normality, homogeneity of covariance matrices, and low multicollinearity among predictors.

##### Normality

Results in **Table 3** indicate significant skewness but non-significant kurtosis effects. While asymmetry was statistically associated with group differences, deviations were not sufficient to invalidate LDA application given the large sample size.

##### Equality of covariance matrices

Box’s M test results in **Table 4** show significant differences in covariance matrices across blood pressure groups (p < 0.05), indicating violation of strict homogeneity. However, prior methodological studies suggest LDA remains robust under moderate violations when sample sizes are large.

##### Multicollinearity

Correlation coefficients in **Table 5** were low across predictors, indicating minimal linear dependency. Variance inflation factor values in **Table 6** were all below conservative thresholds, confirming absence of problematic multicollinearity.

### 3.2 Discriminant analysis results

#### 3.2.1 Discriminant functions

**Table 7** presents coefficients of the discriminant functions. The first function accounted for over 99% of between-group variance and was primarily defined by body temperature and weight, while age and heart rate contributed minimally. The second function explained only a small proportion of remaining variance.

Standardized coefficients in **Table 8** confirm weight as the dominant positive contributor to Function 1, followed by age. Temperature loaded negatively, and heart rate had negligible influence. Function 2 was influenced mainly by age and temperature.

The structural matrix in **Table 9** shows the strongest correlation between weight and Function 1, reinforcing its central role in-group separation. Temperature was most strongly associated with Function 2. Age contributed moderately to both functions.

Classification function coefficients (**Table 10)** indicate that temperature and weight were the most influential predictors across all blood pressure groups, while age and heart rate played secondary roles.

Group centroid values in **Table 11** demonstrate clear separation along the first discriminant function, with hypotensive cases showing negative scores and hypertensive cases positive scores. The second function provided limited additional separation.

Tests of equality of means **(Table 12)** revealed significant differences across blood pressure groups for age, temperature, and weight (p < 0.001), while heart rate differences were not statistically meaningful.

Eigenvalues and canonical correlations in **Table 13** confirm the dominance of the first discriminant function, which captured nearly all variability between groups.

Wilks’ Lambda statistics in Table 14 show that the discriminant functions significantly differentiated blood pressure categories (p < 0.001), with Function 1 accounting for the vast majority of discriminatory power.

#### 3.2.2 Model classification performance

The scatter plot of discriminant scores **(Figure 2)** shows strong separation between blood pressure groups along the first discriminant function. Minimal overlap was observed, particularly for normotensive cases.

**Figure 2:**
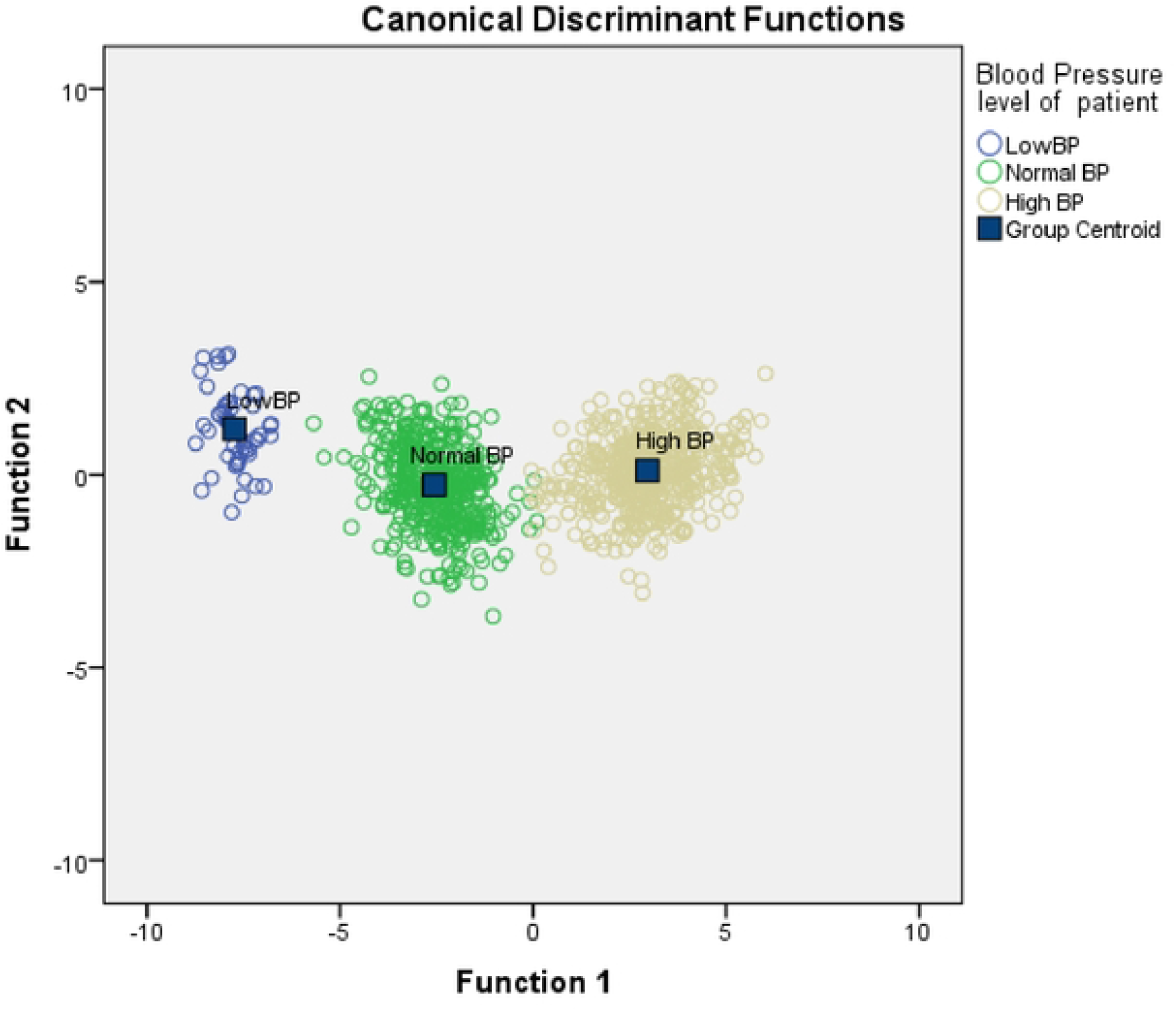
Scatter of Canonical Discriminant Function.

The classification matrix in **Table 15** indicates overall model accuracy of 99.1%. All hypotensive cases were correctly classified. Normotensive and hypertensive groups achieved classification accuracies above 98%. Cross-validation produced identical results, confirming model stability and predictive reliability.

#### 3.2.3 Receiver Operating Characteristic analysis

ROC curves **(Figure 3)** illustrate the classification performance of individual predictors. **Table 16** summarizes area under the curve (AUC) values. Weight demonstrated near-perfect discrimination (AUC = 0.999), and age showed excellent predictive ability (AUC = 0.978). Heart rate and temperature exhibited limited standalone classification power. These findings indicate that weight and age are the strongest individual predictors of blood pressure status in this dataset.

**Figure 3:**
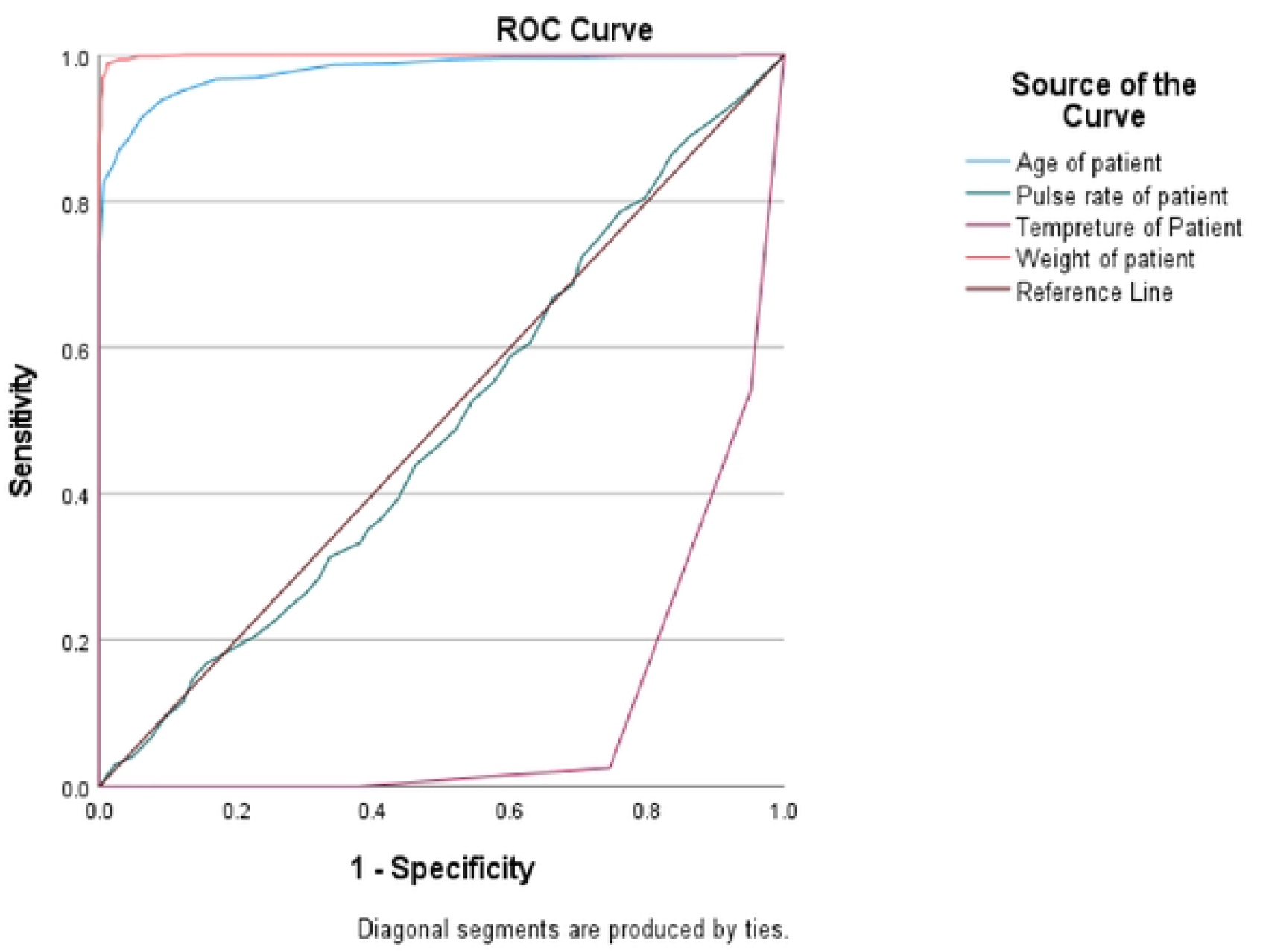
ROC Curve.

## 4. DISCUSSION

### 4.1 Key findings

This study demonstrates that routine vital indicators can accurately classify systolic blood pressure status using discriminant analysis. Body weight emerged as the dominant predictor, accounting for most of the variability between hypotensive, normotensive, and hypertensive groups. Age also contributed meaningfully to classification, while temperature played a secondary but notable role. Heart rate showed minimal discriminative influence. The model achieved extremely high classification accuracy, and cross-validation confirmed its stability, indicating that a small set of routinely collected physiological indicators can effectively differentiate blood pressure categories.

### 4.2 Comparison with literature

The strong influence of body weight aligns with extensive epidemiological and physiological evidence linking excess adiposity to hypertension risk (Janssen et al., 2004; Neter et al., 2003). Increased body mass is associated with elevated peripheral resistance, neurohormonal activation, and vascular remodeling, all of which contribute to sustained blood pressure elevation (Hall et al., 2015). The prominent role of age in the model is consistent with established findings that hypertension prevalence rises with advancing age due to arterial stiffening and cumulative cardiovascular stress (Dorans et al., 2018; Wang & Wang, 2004).

Temperature also showed meaningful association within the discriminant framework. Prior studies report that environmental temperature influences vascular tone and blood pressure regulation, with cold exposure often associated with elevated cardiovascular stress (Cabrera et al., 2016; Lewington et al., 2012). Although temperature demonstrated weaker independent predictive power in ROC analysis, its contribution within the multivariate model supports emerging evidence that thermal stress may act as a contextual cardiovascular modifier.

The high AUC values observed for weight and age confirm their strong standalone predictive ability. This is consistent with pathophysiological models linking obesity and aging to hypertension development (Kotsis et al., 2010). In contrast, the limited contribution of heart rate suggests that resting pulse alone may not sufficiently capture underlying vascular risk in this population.

### 4.3 Implications

These findings have practical implications for clinical and public health screening. Weight and age are easily measurable indicators that can support rapid risk stratification in both hospital and community settings. Incorporating predictive models based on routine vital signs into electronic health systems or mobile health platforms could enable early identification of individuals at elevated risk. Such tools may assist clinicians in prioritizing preventive counseling, lifestyle interventions, and monitoring strategies.

From a public health perspective, the strong association between weight and blood pressure reinforces the importance of obesity prevention and weight management as central strategies in hypertension control. Community-based programs promoting physical activity, dietary modification, and awareness of cardiovascular risk factors may complement clinical interventions and reduce population-level disease burden.

### 4.4 Limitations

The retrospective design restricts causal interpretation and does not account for potential confounding variables such as medication use, socioeconomic status, or comorbid conditions. Additionally, while discriminant analysis demonstrated high accuracy, external validation in independent cohorts is required before clinical implementation.

### 4.5 Conclusion

A discriminant model based on routine vital indicators can classify blood pressure status with high accuracy. Body weight and age are the primary drivers of discrimination, with temperature providing additional contextual contribution. The findings support the feasibility of using simple physiological measurements for scalable hypertension risk stratification. With further validation, this approach could inform targeted prevention strategies and strengthen early detection efforts in both clinical and public health settings.

## Data Availability

The data is available upon request

## AUTHOR STATEMENTS

### Ethical approval

Ethical clearance for this study was obtained from the Ethical Review Committee of Takoradi Technical University prior to data access. The study used anonymized secondary hospital records obtained with institutional permission from the regional hospital. No personally identifiable information was accessed. According to local institutional guidelines, the study involved retrospective analysis of de-identified routine clinical data collected for administrative purposes.

### Funding

This research received no external funding.

### Competing interests

The authors declare no competing interests.

## Notes

### Competing Interest Statement

The authors have declared no competing interest.

### Funding Statement

The author(s) received no specific funding for this work.

